# Model-based evaluation of school- and non-school-related measures to control the COVID-19 pandemic

**DOI:** 10.1101/2020.12.07.20245506

**Authors:** Ganna Rozhnova, Christiaan H. van Dorp, Patricia Bruijning-Verhagen, Martin C.J. Bootsma, Janneke H.H.M. van de Wijgert, Marc J.M. Bonten, Mirjam E. Kretzschmar

**Affiliations:** Julius Center for Health Sciences and Primary Care, University Medical Center Utrecht, Utrecht University, Utrecht, The Netherlands; BioISI—Biosystems & Integrative Sciences Institute, Faculdade de Ciências, Universidade de Lisboa, Lisboa, Portugal; Theoretical Biology and Biophysics (T-6), Los Alamos National Laboratory, Los Alamos, New Mexico, USA; Mathematical Institute, Utrecht University, Utrecht, The Netherlands; The Institute of Infection, Veterinary and Ecological Sciences, University of Liverpool, Liverpool, UK; Department of Medical Microbiology, University Medical Center Utrecht, Utrecht University, The Netherlands

## Abstract

**Background:** In autumn 2020, many countries, including the Netherlands, are experiencing a second wave of the COVID-19 pandemic. Health policymakers are struggling with choosing the right mix of measures to keep the COVID-19 case numbers under control, but still allow a minimum of social and economic activity. The priority to keep schools open is high, but the role of school-based contacts in the epidemiology of SARS-CoV-2 is incompletely understood. We used a transmission model to estimate the impact of school contacts on transmission of SARS-CoV-2 and to assess the effects of school-based measures, including school closure, on controlling the pandemic at different time points during the pandemic.

**Methods and Findings:** The age-structured model was fitted to age-specific seroprevalence and hospital admission data from the Netherlands during spring 2020. Compared to adults older than 60 years, the estimated susceptibility was 23% (95%CrI 20—28%) for children aged 0 to 20 years and 61% (95%CrI 50%—72%) for the age group of 20 to 60 years. The time points considered in the analyses were (i) August 2020 when the effective reproduction number (*R*_*e*_) was estimated to be 1.31 (95%CrI 1.15—2.07), schools just opened after the summer holidays and measures were reinforced with the aim to reduce *R*_*e*_ to a value below 1, and (ii) November 2020 when measures had reduced *R*_*e*_ to 1.00 (95%CrI 0.94—1.33). In this period schools remained open. Our model predicts that keeping schools closed after the summer holidays, in the absence of other measures, would have reduced *R*_*e*_ by 10% (from 1.31 to 1.18 (95%CrI 1.04—1.83)) and thus would not have prevented the second wave in autumn 2020. Reducing non-school-based contacts in August 2020 to the level observed during the first wave of the pandemic would have reduced *R*_*e*_ to 0.83 (95%CrI 0.75—1.10). Yet, this reduction was not achieved and the observed *R*_*e*_ in November was 1.00. Our model predicts that closing schools in November 2020 could reduce *R*_*e*_ from the observed value of 1.00 to 0.84 (95%CrI 0.81—0.90), with unchanged non-school based contacts. Reductions in *R*_*e*_ due to closing schools in November 2020 were 8% for 10 to 20 years old children, 5% for 5 to 10 years old children and negligible for 0 to 5 years old children.

**Conclusions:** The impact of measures reducing school-based contacts, including school closure, depends on the remaining opportunities to reduce non-school-based contacts. If opportunities to reduce *R*_*e*_ with non-school-based measures are exhausted or undesired and *R*_*e*_ is still close to 1, the additional benefit of school-based measures may be considerable, particularly among the older school children.

## Introduction

In autumn 2020, many countries, including the Netherlands, are experiencing a second wave of the COVID-19 pandemic [1]. During the first wave in spring 2020, general population-based control physical distancing measures were introduced in the Netherlands, which included refraining from hand-shaking, work from home if possible, self-isolation of persons with cold- or flu-like symptoms and closure of all schools. These contact-reduction measures were relaxed starting from May, and the incidence of COVID-19 started to increase again at the end of July [1]. From the end of August onward, contact-reduction measures were reintroduced in a step-wise manner. Schools closed during July and August for summer break, reopened at the end of August, and have remained open until this day (December 7, 2020), with the exception of a one-week autumn break. Some measures were implemented in schools after the summer break to reduce transmission. Students and teachers in secondary schools have to wear masks when not seated at their desks, and students have to keep distance from teachers. A student with cold- or flu-like symptoms has to stay at home.

The step-wise increase in control measures after the summer started with earlier closing times of bars and restaurants, reinforcement of working at home (in September), followed by closure of all bars and restaurants, theaters, cinemas and other cultural meeting places in November and obligatory mask wearing in all public places since December 1. Estimated effective reproduction numbers (*R*_*e*_) were about 1.3 at the end of August and about 1.0 since November 13th [1]. The aim of the implemented measures was to reduce *R*_*e*_ to 0.8. The failure to achieve this might be due to reduced societal acceptance of control measures, and/or due to the lack of schools closure. The role of children and their contacts during school hours in the spread of SARS-CoV-2 is in fact not well understood [2]. In this study, we explored this role with a mathematical model fitted to COVID-19 data from the Netherlands.

Closure of schools is considered an effective strategy to contain an influenza pandemic [3], based on both model calculations and observational studies of the influence of school holidays on the spread of influenza [4, 5]. The reasons for this are the high contact rates in young age groups [6] and the susceptibility of children and young people to the influenza virus. In contrast to influenza, children seem to be less susceptible to SARS-CoV-2 than adults and, based on sparse data, the susceptibility to SARS-CoV-2 increases with age [7, 8].

In the absence of empirical SARS-CoV-2 data, mathematical modeling can help to quantify the role of different age groups in the distribution of SARS-CoV-2 in the population [9], and to evaluate the impact of interventions on transmission [10–13]. Such models can estimate the reduction in the effective reproduction number for different contact-reduction scenarios within or outside school environments. Model predictions about the relative epidemic impacts of school- and non-school-based measures can assist policymakers to select sets of measures during different stages of the pandemic that optimally balance potential harms and benefits. Predictions generated by models that include differences in susceptibility and contract rates in different age groups can also aid in deciding which school age groups should be the primary target of school-based interventions.

We used an age-structured transmission model for SARS-CoV-2 fitted to the number of hospital admissions due to COVID-19 and seroprevalence during spring 2020 in the Netherlands to evaluate the impact of reducing school and other (non-school-related) contacts in society to control the second wave of COVID-19 in the autumn of 2020. We provide a comparative impact of these measures on the effective reproduction number in August 2020, before the most recent set of measures was implemented, and in November 2020, when the most recent measures were still in place. We assess which combinations of school and non-school related measures are most likely successful in reducing the reproduction number to below 1 and which school ages should be targeted to design effective school-based interventions.

## Methods

### Overview

Estimates of epidemiological parameters were obtained by fitting a transmission model to age-stratified COVID-19 hospital admission data (*n* = 10, 961) and cross-sectional age-stratified SARS-CoV-2 seroprevalence data (*n* = 3, 207) [14]. The model equipped with parameter estimates was subsequently used to investigate the impact of school- and non-school-based measures on controlling the pandemic.

### Data

The hospital data included *n* = 10, 961 COVID-19 hospitalizations by date of admission and stratified by age during the period of 69 days following the first official case in the Netherlands (27 February 2020).

The SARS-CoV-2 seroprevalence data was taken from a cross-sectional population-based serological study carried out in April-May 2020 [14]. A total of 40 municipalities were randomly selected from the Netherlands, with probabilities proportional to their population size. From these municipalities, an age-stratified sample was drawn from the population register, and 6, 102 persons were invited to participate. Serum samples and questionnaires were obtained from 3, 207 participants and included in the analyses. The majority of blood samples were drawn in the first week of April.

Our analyses made use of the demographic composition of the Dutch population in July 2020 from Statistics Netherlands [15] and age-stratified contact data for the Netherlands [16, 17]. The contact rates before the pandemic were based on a cross-sectional survey carried out in 2016/2017, where participants reported the number and age of their contacts during the previous day [16]. The contact rates after the first lockdown were based on the same survey which was repeated in a sub-sample of the participants in April 2020 (PIENTER Corona study) [16]. School-specific contact rates for the Dutch population before the pandemic were taken from the POLYMOD study [6, 17].

### Transmission model

We used a deterministic compartmental model describing SARS-CoV-2 transmission in a population stratified by infection status and age (Figure 1 A). The dynamics of the model follows the Susceptible-Exposed-Infectious-Recovered structure. Persons in age group *k*, where *k* = 1, …, *n*, are classified as susceptible (*S*_*k*_), infected but not yet infectious (exposed, *E*_*k*_), infectious in *m* stages (*I*_*k,p*_, where *p* = 1, …, *m*), hospitalized (*H*_*k*_) and recovered without hospitalization (*R*_*k*_). Susceptible persons (*S*_*k*_) can acquire infection via contact with infectious persons (*I*_*k,p*_) and become latently infected (*E*_*k*_) at a rate *β*_*k*_*λ*_*k*_, where *λ*_*k*_ is the force of infection, and *β*_*k*_ is the reduction in susceptibility to infection of persons in age group *k* compared to persons in age group *n*. After the latent period (duration 1*/α* days), exposed persons become infectious (*I*_*k*,1_). Infectious persons progress through (*m* − 1) stages of infection (*I*_*k,p*_, where *p* = 2, …, *m*) at rate *γm*, after which they recover (*R*_*k*_). Inclusion of *m* identical infectious stages allows for the tuning of the distribution of the infectious period, interpolating between an exponentially distributed infectious period (*m* = 1) and a fixed infectious period (*m* → ∞). Intermediate values of *m* correspond to an Erlang-distributed infectious period with mean 1*/γ* and standard deviation 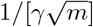. Hospitalization (*H*_*k*_) of infectious persons (*I*_*k,p*_) occurs at rate *v*_*k*_. Since the model is fit to hospital admissions data, the disease-related mortality and discharge from the hospital are not explicitly modeled. Given the timescale of the pandemic and the lack of reliable data on reinfections, we assume that recovered individuals cannot be reinfected. As the timescale of the pandemic is short compared to the average lifespan of persons, we neglected natural birth and death processes, and the population size in the model stays constant.

**Figure 1.**
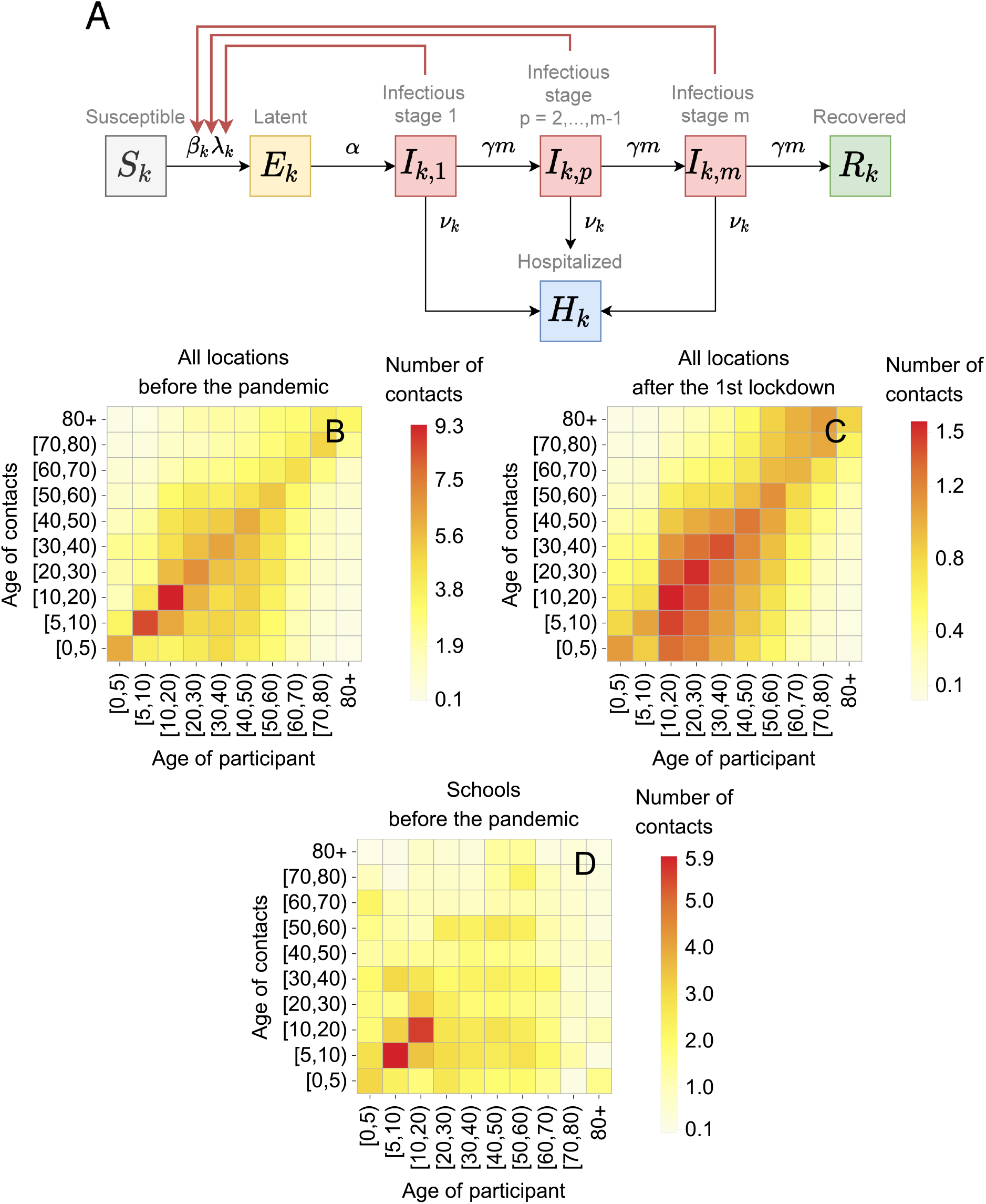
(A) Schematic of the transmission model. Black arrows show epidemiological transitions. Red arrows indicate the compartments contributing to the force of infection. Susceptible persons in age group *k* (*S*_*k*_), where *k* = 1, …, *n*, become latently infected (*E*_*k*_) via contact with infectious persons in *m* infectious stages (*I*_*k,p*_, *p* = 1, …, *m*) at a rate *β*_*k*_*λ*_*k*_, where *λ*_*k*_ is the force of infection, and *β*_*k*_ is the reduction in susceptibility to infection of persons in age group *k* compared to persons in age group *n*. Exposed persons (*E*_*k*_) become infectious (*I*_*k*,1_) at rate *α*. Infectious persons progress through (*m* − 1) infectious stages at rate *γm*, after which they recover (*R*_*k*_). From each stage, infectious persons are hospitalized at rate *v*_*k*_. Table 1 gives the summary of the model parameters. **(B)-(D) Contact rates**. (B) and (C) show contact rates in all locations before the pandemic and after the first lockdown (April 2020), respectively. (D) shows contact rates at schools before the pandemic. The color represents the average number of contacts a person in a given age group had with persons in another age group.

We assume that, before the first lockdown, the probability of transmission per contact between a susceptible and an infectious individual, *E*, is independent of the age of two individuals. After introduction of the control measures in March 2020, this probability of transmission decreased to *Eζ*_1_, where 0 ≤ *ζ*_1_ ≤ 1. (1 − *ζ*_1_) then denotes the reduction in the probability of transmission due to general population-based measures that are not explicitly included in the model, such as refraining from shaking hands, physical distancing, mask-wearing, and self-isolation of symptomatic persons. We denote the general contact rate of a person in age group *k* with persons in age group *l, c*_*kl*_, and the contact rates specific to the periods before and after the first lockdown, *b*_*kl*_, and, *a*_*kl*_, respectively (see Figures 1 B and C). We model the transition in the general contact rate using a linear combination

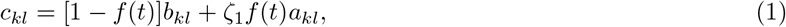

where the contribution of the contact rate after the first lockdown is given by the logistic function

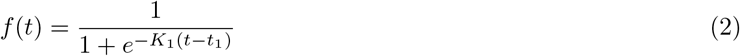

with the mid-point value *t*_1_ and the logistic growth *K*_1_. The parameter *K*_1_ governs the speed at which control measures are rolled out, and *t*_1_ is the mid-time point of the lockdown period (Figure S1). The special cases of *f* = 0 and *f* = 1 describe the contact rate before and after the first lockdown, with *f* values between 0 and 1 corresponding to contact rates at the intermediate time points.

**Table 1.** Summary of the model parameters.

| Description (unit) | Notation | Reference |
| --- | --- | --- |
| **Constant parameters** |  |  |
| Number of age groups | $n$ | 10 |
| Number of infectious stages | $m$ | 3 |
| Basic reproduction number | $R_0$ | Estimated using the method in [18] |
| Effective reproduction number | $R_e$ | Estimated using the method in [18] |
| Probability of transmission per contact | $\epsilon$ | Estimated |
| Reduction in post-lockdown probability of transmission per contact | $(1 - \zeta_1)$ | Estimated |
| Latent period (days) | $1/\alpha$ | Estimated |
| Rate of moving between infectious stages (1/day) | $\gamma m$ | Estimated |
| Contribution of the contact rate after the lockdown | $f(t) = 1/[1 + e^{-K_1(t-t_1)}]$ | Eq. 2 |
| Mid-point value of the logistic function (days) | $t_1$ | Estimated |
| Logistic growth (1/day) | $K_1$ | Estimated |
| Over-dispersion parameter for the NegBinom distribution for hospitalizations | $r$ | Estimated |
| Proportion of school contacts | $\omega$ | [0,1] |
| Reduction in probability of transmission per contact during relaxation | $(1 - \zeta_2)$ | [0,1], $\zeta_2 \geq \zeta_1$ |
| Initial fraction of infected persons | $\theta$ | Estimated |
| **Age-specific parameters\*** |  |  |
| Force of infection (1/day) | $\lambda_k$ | Eq. 5 |
| Hospitalization rate (1/day) | $\nu_k$ | Estimated |
| Susceptibility of age group $k$ relative to age group $n$ | $\beta_k$ | Estimated |
| General contact rate (1/day) | $c_{kl}$ | Eqs. 1 and 3 |
| Contact rate before the pandemic (1/day) | $b_{kl}$ | [16] |
| Contact rate after the first lockdown (1/day) | $a_{kl}$ | [16] |
| School contact rate before the pandemic (1/day) | $s_{kl}$ | [6, 17] |
| Population size of age group $k$ | $N_k$ | [15] |

\*Indices $k$ and $l$ denote the age groups $k, l = 1, \dots, n$ .

Similarly, the contact rate incorporating the relaxation of control measures after the first lockdown is modeled as follows

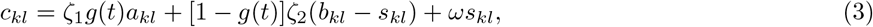

where 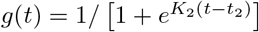 with the mid-point value *t*_2_ *> t*_1_ and the logistic growth *K*_2_. In Eq. 3, the first two terms describe the increase of non-school contacts from the level after the first lockdown to their pre-lockdown level. The parameter *ζ*_2_ ≥ *ζ*_1_, 0 ≤ *ζ*_2_ ≤ 1 implies that the probability of transmission increased due to reduced adherence to control measures. The last term describes opening of schools which we assume to happen instantaneously, where *s*_*kl*_ denotes the school contact rate at the pre-lockdown level (Figure 1 D), and *ω*, 0 ≤ *ω* ≤ 1 is the proportion of retained school contacts. Schools functioning without any measures correspond to *ω* = 1. Schools closure is achieved by setting *ω* = 0. The summary of the model parameters is given in Table 1.

### Model equations

The model was implemented using a system of ordinary differential equations as follows

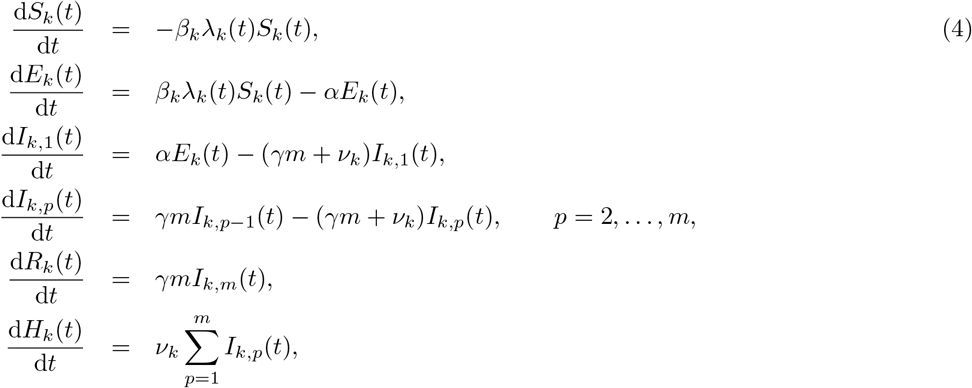

where *S*_*k*_, *E*_*k*_, *R*_*k*_ and *H*_*k*_ are the numbers of persons in age group *k, k* = 1, …, *n*, who are susceptible, exposed, recovered and hospitalized, respectively. The number of infectious persons in age group *k* and stage *p* = 1, …, *m* is denoted *I*_*k,p*_. The force of infection is given by

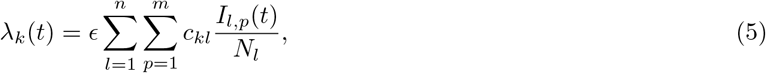

where *N*_*k*_ is the number of individuals in age group 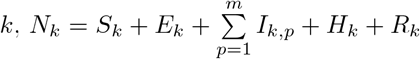. We took 22 February 2020 as starting date (*t*_0_) for the pandemic in the Netherlands, which is 5 days prior to the first officially notified case. We assumed that there were no hospitalizations during this 5 day period. As initial condition for the model, we assume that a fraction *θ* of each age group was infected at time *t*_0_, equally distributed between the exposed and infectious persons, i.e., 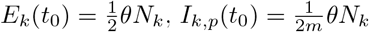 and *S*_*k*_(*t*_0_) = (1 − *θ*)*N*_*k*_.

The model was implemented in Mathematica 10.0.2.0. The code reproducing the results of this study is available at https://github.com/lynxgav/COVID19-schools.

### Observation model and parameter estimation

Given predictions of the model, we calculated the likelihood of the data as follows. In the model, infectious individuals are hospitalized at a continuous rate 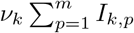. However, the hospitalization data consists of a discrete number of hospital admissions *h*_*k,i*_ on day *T*_*i*_ for each age class *k*. As the probability of hospitalization is relatively small, we made the simplifying assumption that the daily incidence of hospitalizations is proportional to the prevalence of infectious individuals at that time point. To accommodate errors in reporting and within age class variability of the hospitalization rate, we allowed for over-dispersion in the number of hospitalizations using a Negative-Binomial distribution, i.e.,

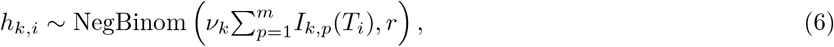

where we parameterize the NegBinom(*µ, r*) distribution with the mean *µ* and over-dispersion parameter *r*, such that the variance is equal to *µ* + *µ*^2^*/r*.

We calculated the likelihood of the seroprevalence data using the model prediction of the fraction of non-susceptible individuals in each age class 1 − *S*_*k*_(*T*)*/N*_*k*_. Here *T* denotes the median sampling time minus the expected duration from infection to seroconversion. We assumed that the probability of finding a seropositive individual in a random sample from the population is equal to the fraction of non-susceptible individuals, leading to a Binomial distribution for the number of positive samples *C*_*k*_ among all samples *L*_*k*_ from age group *k*

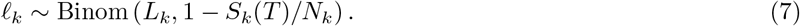

Parameters were estimated in a Bayesian framework using methods we developed before [19, 20]. We used age-specific contact rates with ten age groups, defined by the following age intervals [0,5), [5,10), [10,20), [20,30), [30,40), [40,50), [50,60), [60,70), [70,80) years old and the group of all persons older than 80 years referred to as 80+ age group. Due to the low number of hospitalizations in young persons, we assumed that hospitalization rates in the first three age groups (i.e., [0,20) years old) were equal. The relative susceptibility was estimated for persons in [0,20), [20,60) and 60+ age categories, where 60+ age category was used as the reference [7]. As the age groups for which the seroprevalence was reported [14] are different from the age groups used in our model, we used demographic data from the Netherlands [15] and the smoothed age-specific seroprevalence curve estimated by Vos *et al*. [14] to correct for this discrepancy. The Bayesian prior distributions for the estimated parameters (see Table 1) are listed in Table 2. In the main text, we presented results for three infectious classes (*m* = 3) corresponding to Erlang-distributed infectious periods. The model was fitted to the data using the Hamiltonian Monte Carlo method as implemented in Stan (https://www.mc-stan.org) [21]. We used 4 parallel chains of length 1500 with a warm-up phase of length 1000, resulting in 2000 parameter samples from the posterior distribution. The data and the Stan and R scripts with all parameter settings are available at https://github.com/lynxgav/COVID19-schools.

**Table 2.** Prior distributions for the Bayesian statistical model. The scale parameter of the normal and log-normal distributions is equal to the standard deviation.

| Parameter | Prior | Explanation |
| --- | --- | --- |
| $\epsilon$ | Uniform(0, 1) | flat prior |
| $\alpha$ | InvGamma(32.25, 9.75) | 99% of the prior density of $1/\alpha$ between 2 and 5 days |
| $\gamma$ | InvGamma(22.6, 2.44) | 99% of the prior density of $1/\gamma$ between 5 and 15 days |
| $\nu_k$ | folded-$\mathcal{N}(0, 5)$ | vague prior |
| $\beta_{[0,20)}$ | LogNormal(-1.47, 0.1) | Log-odds $-1.47 = \log(0.23)$ based on prior estimates [7] |
| $\beta_{[20,60)}$ | LogNormal(-0.45, 0.1) | Log-odds $-0.45 = \log(0.64)$ based on prior estimates [7] |
| $r$ | LogNormal(5, 2) | vague prior |
| $\zeta_1$ | folded-$\mathcal{N}(1, 0.1)$ | *a priori*, we expect the reduction in contacts after the first lockdown to account for most of the decrease in the transmission rate |
| $t_1$ | $\mathcal{N}(23, 7)$ | the mean of $t_1$ is given by the day of initiation of most drastic social distancing measures (March 15); most measures were taken within two weeks from this date |
| $K_1$ | Exp(1) | with $K_1 = 1$ the uptake of measures takes approximately 6 days |
| $\theta$ | Uniform($10^{-7}, 5 \cdot 10^{-4}$) | vague prior allowing for approximately $10^0$–$10^5$ infections at time $t_0$ |

### Model outcomes

We considered control measures aimed at reducing contact rate at schools or in all other locations. Main outcome measures were age-specific seroprevalence and hospital admissions. In addition, we evaluated the impact of a control measure by computing the effective reproduction number (*R*_*e*_) using the next generation matrix method [18, 22], and percentage of contacts that need to be reduced to achieve control of the pandemic as quantified by *R*_*e*_ = 1.

## Results

### Epidemic dynamics

The model shows a very good agreement between the estimated age-specific hospitalizations and the data (Figure 2). The number of hospitalizations increases with age, with the highest peaks in hospitalizations observed in persons above 60 years old. The estimated probability of hospitalization increases nearly exponentially with age (as shown by an approximately linear relationship on the logarithmic scale, Figure 3), except for persons under 30 years old, in whom the number of hospitalizations was low. The estimated probability of hospitalization increased from 0.09% (95%CrI 0.05—0.15%) in persons under 20 years old to 4.37% (95%CrI 2.80—8.82%) in persons older than 80 years (Figure S2).

**Figure 2.**
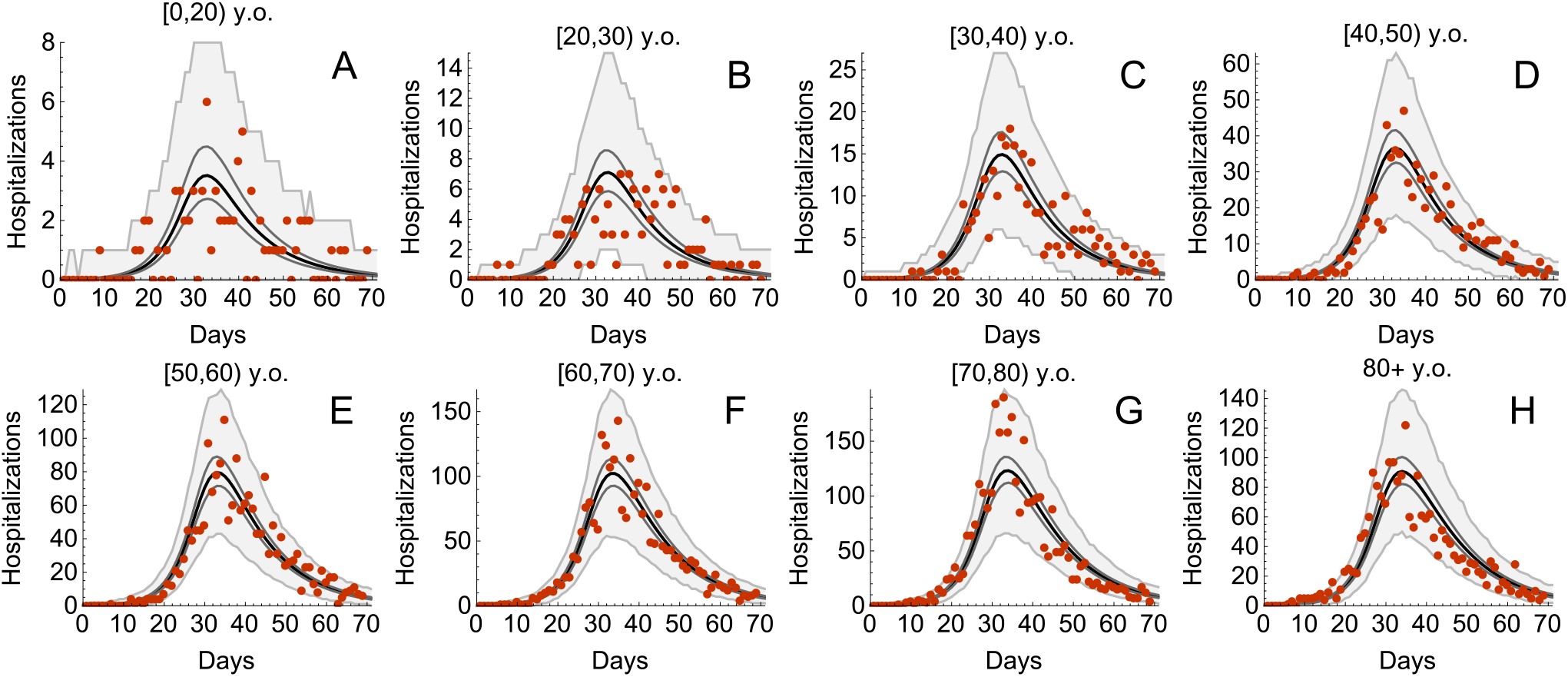
Estimated age-specific hospital admissions. The black lines represent the estimated medians. The dark gray lines correspond to 95% credible intervals obtained from 2000 parameter samples from the posterior distribution, and the shaded region shows 95% Bayesian prediction intervals. The dots are daily hospitalization admission data.

**Figure 3.**
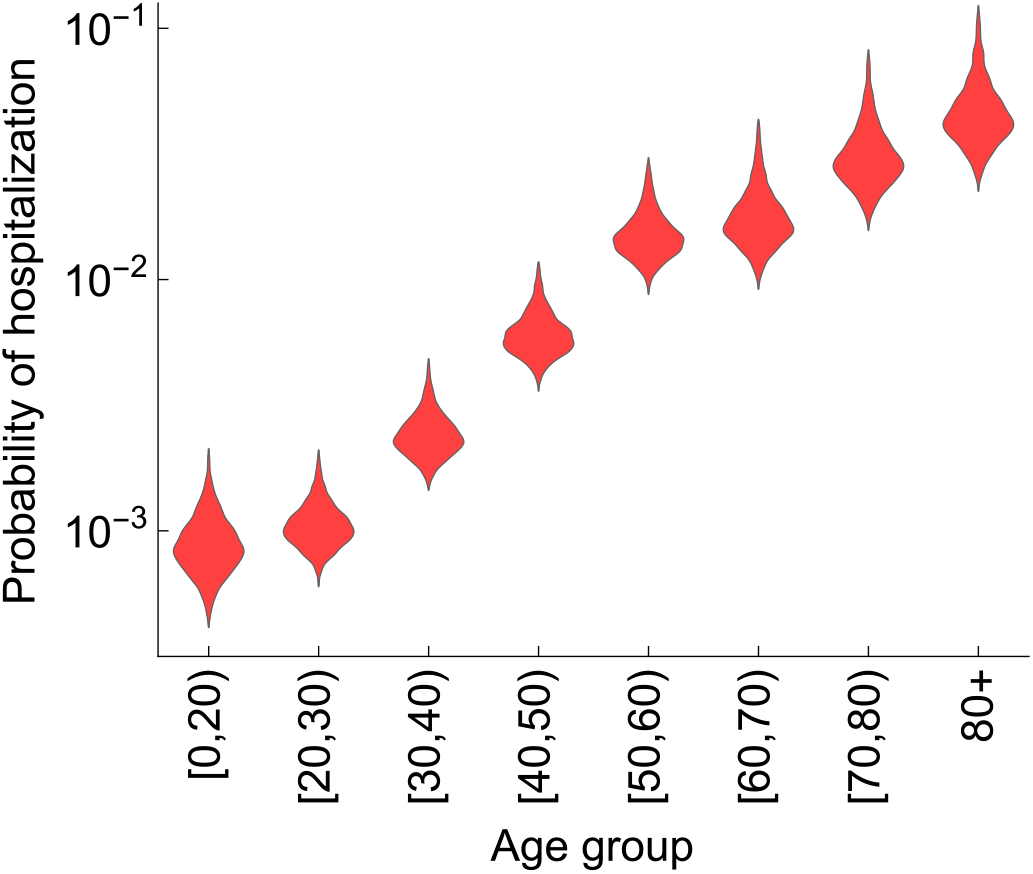
Estimated age-specific probability of hospitalization. The violin shapes represent the marginal posterior distribution of the probability of hospitalization in the model. The y-axis is shown on the log10 scale.

The model accurately reproduces the percentage of seropositive persons distributed across the age groups (Figure 4). The median seroprevalence in the population was 2.7%, with the maximum seroprevalence observed in persons between 20 and 40 years old (about 3.5%). The lowest seroprevalence was among children in the 0 to 10 years age group (0.9%). Note that if our model did not include age-dependence of susceptibility to SARS-CoV-2, the seroprevalence peak would be expected among children because they have the largest numbers of contacts in the population.

**Figure 4.**
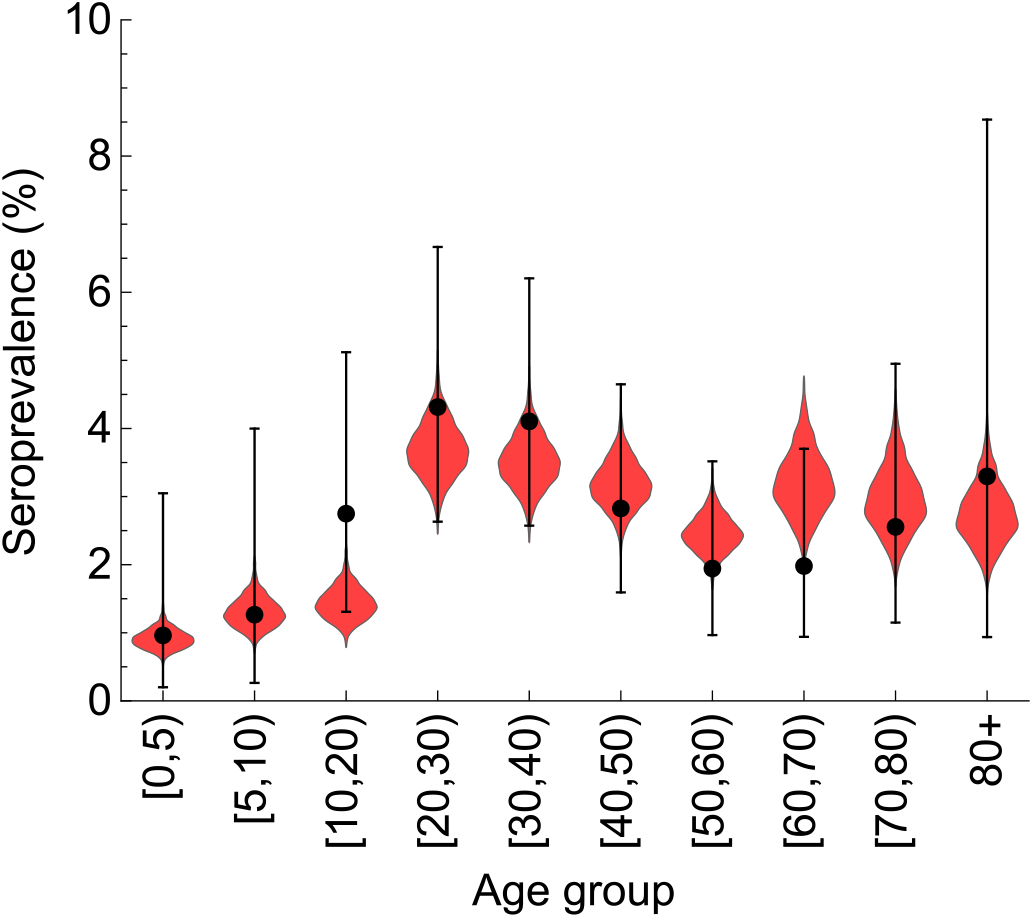
Estimated age-specific seroprevalence. The dots and error bars show the percentage of seropositive persons based on the data. The error bars represent the 95% confidence (Jeffreys) interval of the percentage. The violin shapes represent the marginal posterior distribution of the percentage of seropositive persons in the model.

The estimated probability of transmission per contact was 0.07 (95%CrI 0.05—0.12) before the first lockdown and it decreased by 48.84% (95%CrI 23.81—87.44%) after the first lockdown. The reduction in susceptibility relative to susceptibility in persons above 60 years old was 23% (95%CrI 20—28%) in persons under 20 years old and 61% (95%CrI 50—72%) for persons between 20 and 60 years old (Figure S3). The estimated basic reproduction number was 2.71 (95%CrI 2.15—5.18) in the absence of control measures (February 2020) (Figure S4 A), and dropped to 0.62 (95%CrI 0.29—0.74) after the full lockdown (April 2020) (Figure S4 B). Figures S1, S2, S3, and S4 show an overview of all parameter estimates which are not given in the main text.

### School and non-school based measures

The sequence of measures implemented and lifted during the pandemic in the Netherlands and the respective estimated values of the effective reproduction numbers are shown schematically in Figure 5. We used the fitted model to separately determine the effect on the effective reproduction number of decreasing contacts in schools and of decreasing other (non-school-related) contacts in society in August 2020 (Figure 6) and November 2020 (Figure 7). In doing so, we varied one type of contact and kept the other type constant. For each scenario, the reduction in contact rate was varied between 0% and 100%. The aim of reducing the number of contacts of each type is to decrease the effective reproduction number below 1.

**Figure 5.**
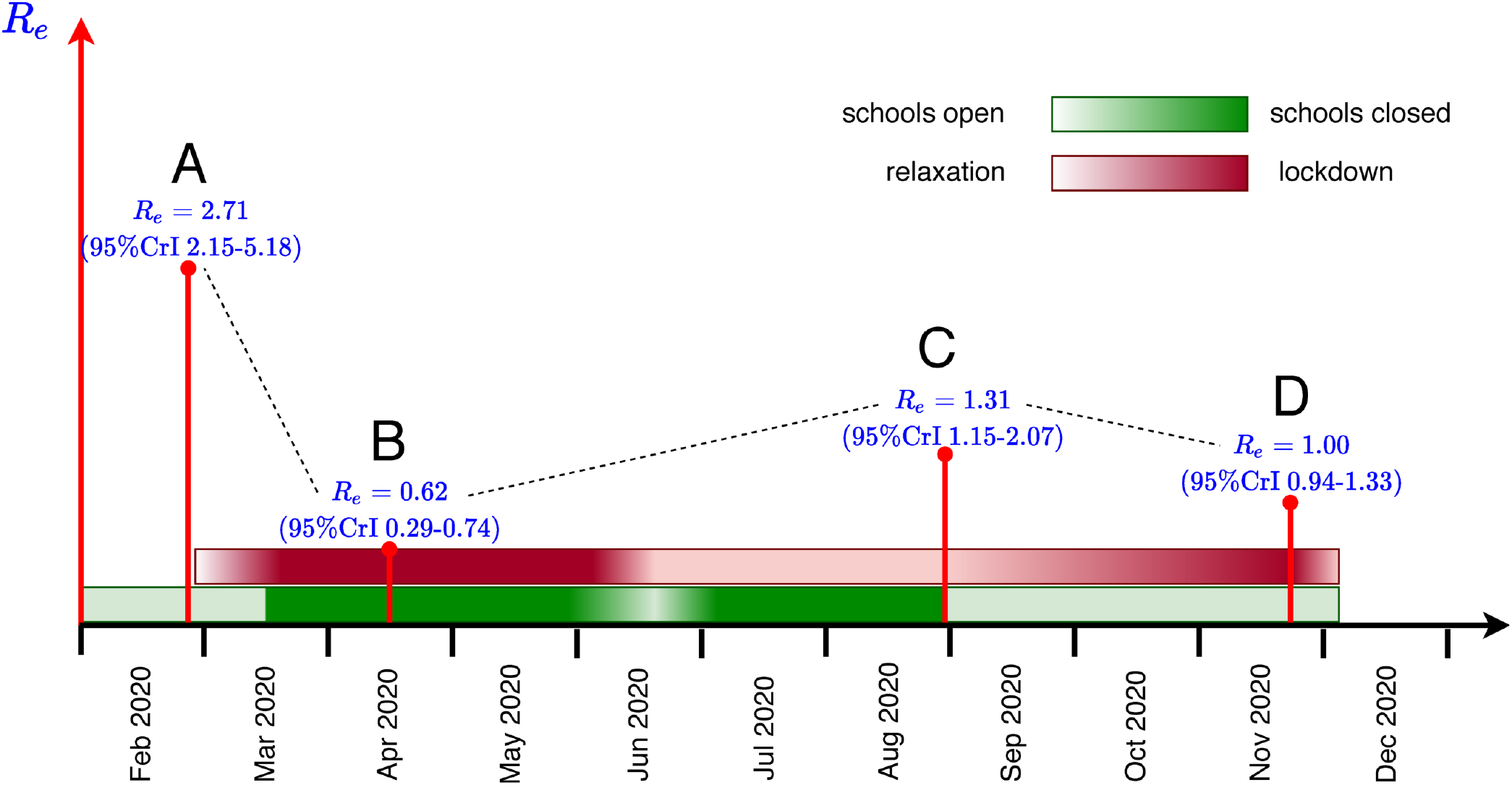
Schematic timeline of the pandemic in the Netherlands. Outlined are times of the introduction and relaxation of control measures, and the estimated effective reproduction numbers for A - start of the pandemic (February 2020), B - full lockdown (April 2020), C - schools opening (August 2020), D - partial lockdown (November 2020). See Figure S4 for the estimated reproduction numbers.

**Figure 6.**
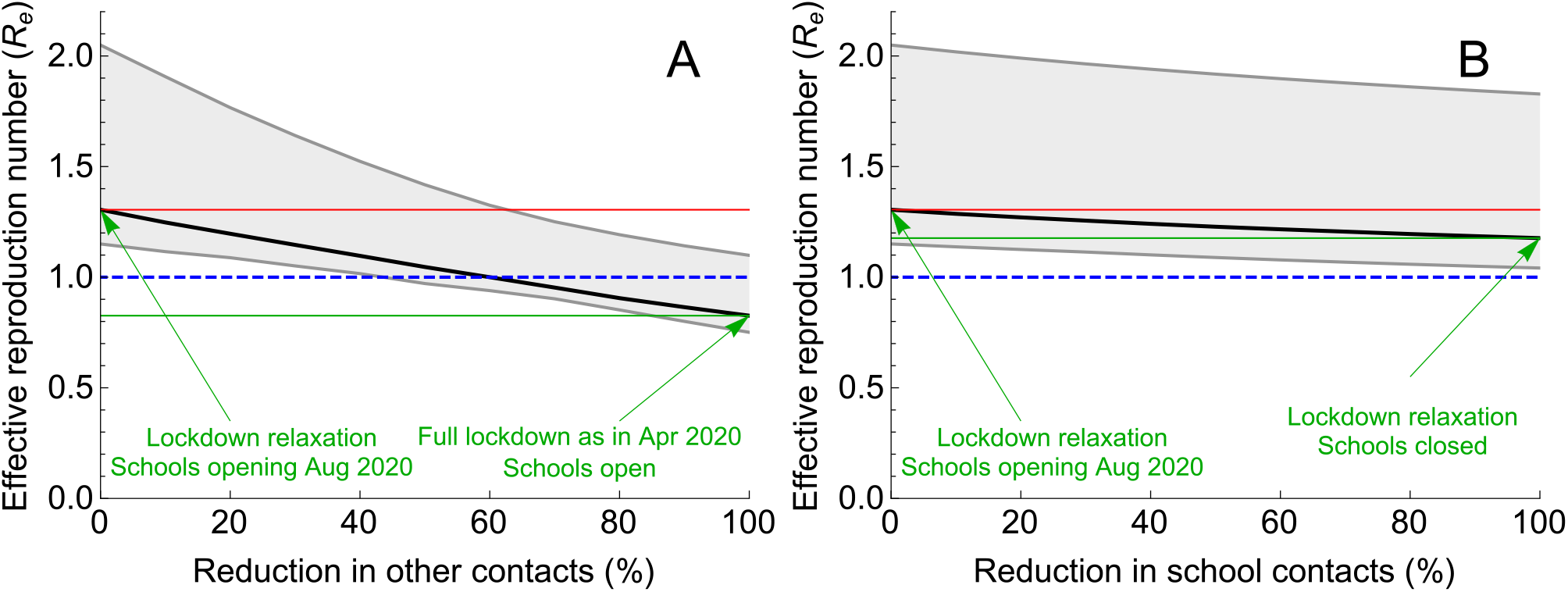
Impact of reduction of two types of contacts on the effective reproduction number in August 2020. Percentage reduction in (A) other (non-school related) contacts and (B) school contacts, with the number of the other type of contact kept constant in each of the two panels. The scenario with 0% reduction describes the situation in August 2020, when schools just opened in the Netherlands. The scenario with 100% reduction represents a scenario with either (A) maximum reduction in other (non-school related) contacts to the level of April 2020 or (B) complete closure of schools. The solid black line describes the median, the shaded region represents the 95% credible intervals obtained from 2000 parameter samples from the posterior distribution. The red line is the starting value of *R*_*e*_ (situation August 2020), the green line is the value of *R*_*e*_ achieved for 100% reduction in contacts. The blue line indicates *R*_*e*_ of 1. To control the pandemic, *R*_*e*_ *<* 1 is necessary.

**Figure 7.**
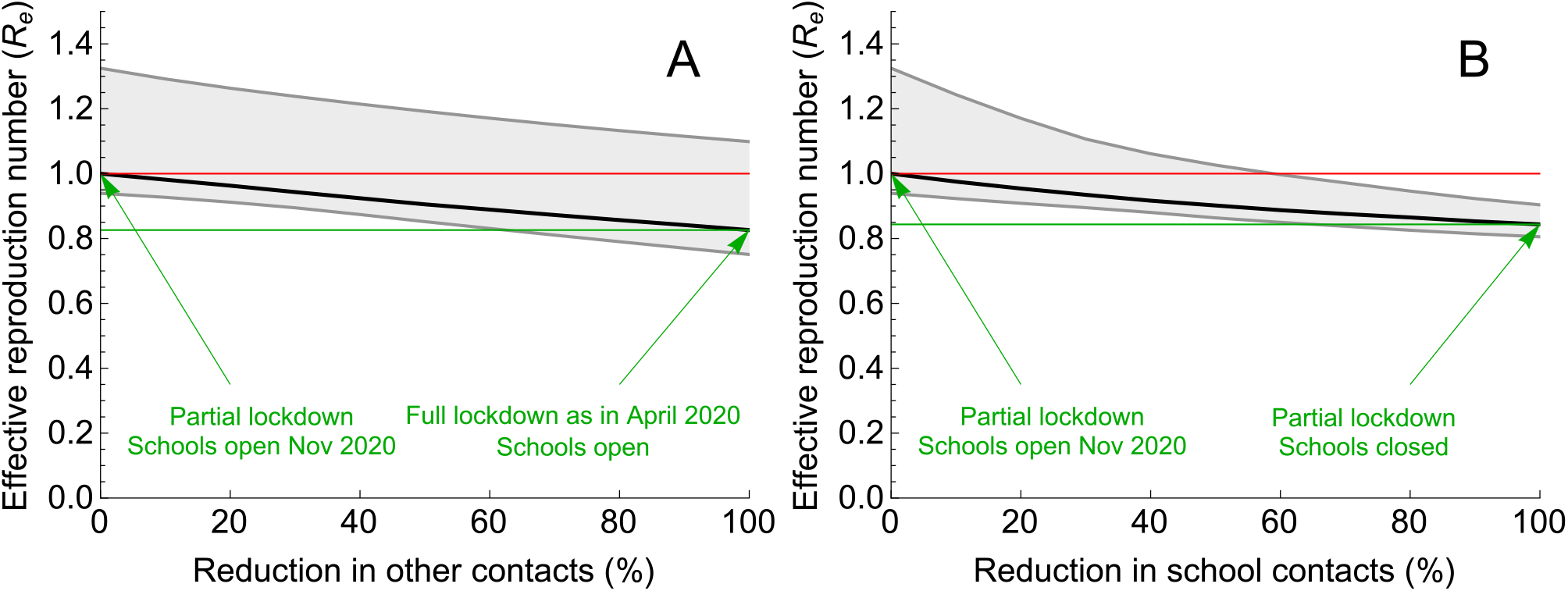
Impact of reduction of two types of contacts on the effective reproduction number in November 2020. Percentage reduction in (A) other (non-school related) contacts and (B) school contacts, with the number of the other type of contact kept constant in each of the two panels. The scenario with 0% reduction describes the situation in November 2020. The scenario with 100% reduction represents a scenario with either (A) maximum reduction in other (non-school related) contacts to the level of April 2020 or (B) complete closure of schools. The solid black line describes the median, the shaded region represents the 95% credible intervals obtained from 2000 parameter samples from the posterior distribution. The red line is the starting value of *R*_*e*_ (situation November 2020), the green line is the value of *R*_*e*_ achieved for 100% reduction in contacts. To control the pandemic, *R*_*e*_ *<* 1 is necessary.

We first considered the situation in August 2020 (Figure 6), when schools had just opened after the summer holidays and when general control measures in the population were less stringent than in April (full lockdown). Between August and December 2020, the only infection prevention measure in primary schools was the advice to teachers and pupils to stay at home in case of symptoms or a household member diagnosed with SARS-CoV-2 infection; physical distancing between teachers and pupils (but not between pupils) only applied to secondary schools. We therefore assumed that the effective number of contacts in schools was the same as before the pandemic (*ω* = 1). For the non-school related contacts we assumed that 1) the number of contacts increased after April 2020 (full lockdown) but was lower than before the pandemic, and that 2) the transmission probability per contact was lower due to general physical distancing and hygiene measures. The starting point of our analyses is an effective reproduction number of 1.31 (95%CrI 1.15—2.07) in accordance with the situation in August 2020 (Figure S4 C). Specifically, to achieve *R*_*e*_ = 1.31 we fixed *ζ*_2_ at 0.67 (decrease in adherence to contact-reduction measures in August as compared to April, when *ζ*_1_ is estimated at 0.51) and *g* at 0.5 (half-way in the relaxation of non-school contacts).

Assuming the state of the Dutch pandemic in August 2020, Figure 6 A demonstrates that non-school related contacts would have to be reduced by at least 50% to bring the effective reproduction number to 1 (if school related contacts do not change). A 100% reduction would resemble the number of contacts in April (full lockdown) and would bring the effective reproduction number to 0.83 (95%CrI 0.75—1.10). Figure 6 B demonstrates that reductions of school contacts would have a limited impact on the effective reproduction number (if non-school contacts do not change). A 100% reduction (complete closure of schools) would have reduced the effective reproduction number by only 10% (from 1.31 to 1.18 (95%CrI 1.04—1.83)).

Subsequently, we considered the Dutch pandemic situation in November 2020 (Figure 7), when the measures implemented since the end of August (partial lockdown intended to prevent the second wave) had led to an effective reproduction number of 1.00 (95%CrI 0.94—1.33) (Figure S4 D), and, as described above, only limited control measures were taken in schools. Now, the impact of interventions targeted at reducing school contacts (Figure 7 B) would reduce the effective reproduction number similarly as reducing non-school contacts in the rest of the population (Figure 7 A). Specifically, closing schools would reduce the effective reproduction number by 16% (from 1.0 to 0.84 (95%CrI 0.81—0.90)) (Figure 7 B). Almost the same *R*_*e*_, i.e., 0.83 (95%CrI 0.75—1.10), would have been achieved by reducing non-school related contacts to the level of April 2020 while the schools remain open (Figure 7 A).

### Interventions for different school ages

Next we investigated the impact of targeting interventions at different age groups, starting from the situation in November 2020 with the effective reproduction number being 1 (Figure S4 D). Figure 8 A, B, and C show *R*_*e*_ as a function of the reduction of school contacts in age groups of [0,5), [5,10) and [10,20) y.o., respectively. In each panel, we varied the number of school contacts in one age group while keeping the number of school contacts in the other two age groups constant. 0% reduction corresponds to the situation in November 2020, and 100% reduction represents a scenario with schools for students in a given age group closed. The model predicts a maximum impact on *R*_*e*_ from reducing contacts of 10 to 20 year old children (Figure 8 C). Closing schools for this age group only could decrease *R*_*e*_ by about 8% (compare Figure 8 C and Figure 7 B where we expect the reduction of 16% after closing schools for all ages). Schools closure for children aged 5 to 10 years would reduce *R*_*e*_ by about 5% (Figure 8 B). Contact reductions among 0 to 5 year old children would have negligible impact on *R*_*e*_ as shown in Figure 8 A.

**Figure 8.**
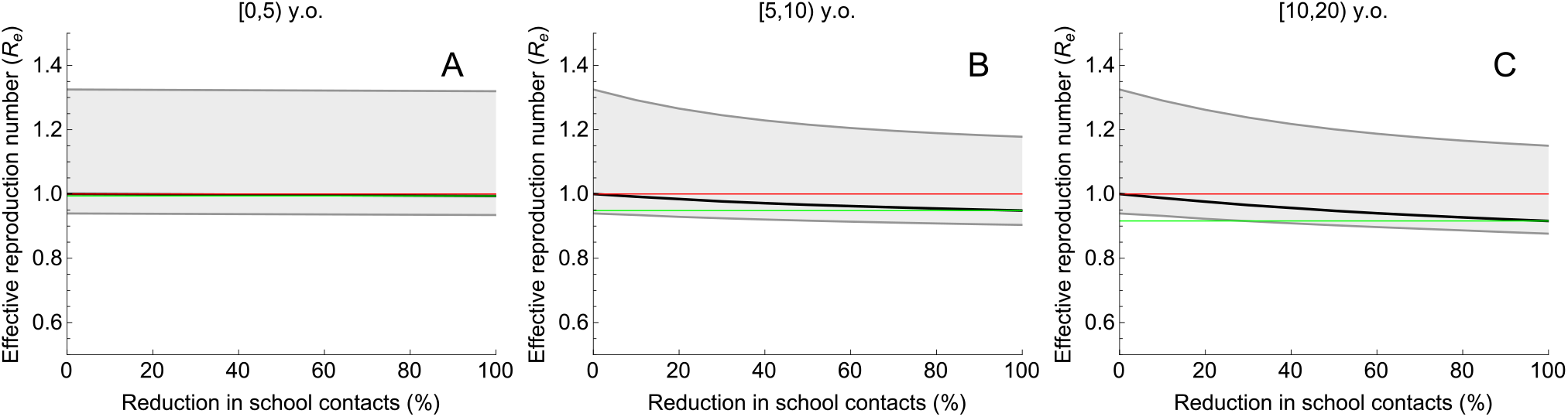
Impact of reduction of school contacts in different age groups on the effective reproduction number in December 2020. Percentage reduction in school contacts among (A) [0, 5) y.o., (B) [5, 10) y.o. and (C) [10, 20) y.o. In each panel, we varied the number of school contacts in one age group while keeping the number of school contacts in the other two age groups constant. The scenario with 0% reduction describes the situation in November 2020 with *R*_*e*_ of about 1 (partial lockdown intended to prevent the second wave), where all schools are open without substantial additional measures. The reduction of 100% in school contacts represents a scenario with the structure of non-school contacts as in November 2020 and schools for students in a given age group closed. The solid black line describes the median, the shaded region represents the 95% credible intervals obtained from 2000 parameter samples from the posterior distribution. The red line is the starting value of *R*_*e*_ = 1 (situation November 2020). The green line indicates the value of *R*_*e*_ achieved when schools for a given age group close.

## Discussion

We used an age-structured model for SARS-CoV-2 fitted to hospital admission and seroprevalence data during spring 2020 to estimate the impact of school contacts on transmission of SARS-CoV-2 and to assess the effects of school-based measures, including schools closure, to mitigate the second wave in the autumn of 2020. We demonstrate how the relative impact of school-based measures aimed at reduction of contacts at schools on the effective reproduction number increases when the effects of non-school-based measures appear to be insufficient. These findings underscore the dilemma for policymakers of choosing between stronger enforcement of population-wide measures to reduce non-school-based contacts or measures that reduce school-based contacts, including complete closure of schools. For the latter choice, our model predicts highest impact from measures implemented for the oldest school ages. We used the Netherlands as a case example but our model code is freely available and can be readily adapted to other countries given the availability of hospitalization and seroprevalence data. The findings in our manuscript can be relevant for guiding policy decisions in the Netherlands, but also in countries where the contact structure in the population is similar to that of the Netherlands [6].

Our model integrates prior knowledge of epidemiological parameters and the quantitative assessment of the model uncertainties in a Bayesian framework. The model has been carefully validated to achieve an excellent fit to data of daily hospitalizations due to COVID-19 and seroprevalence by age. Furthermore, reproduction numbers at different time points of the pandemic correlated well with estimates obtained from independent sources [1]. Finally, susceptibility to infection with SARS-CoV-2 was estimated to increase with age, which corroborates published findings [7,8]. Compared to adults older than 60 years, the estimated susceptibility was about 20% for children aged 0 to 20 years and about 60% for the age group of 20 to 60 years. However, even with extensive validation, we need to be careful when interpreting the predictions of our model as these depend on the sensitivity of serology to identify individuals with prior infection. Recent studies suggest that in persons who experience mild or asymptomatic infections, SARS-CoV-2 antibodies may not always be detectable post-infection [23, 24].

Naturally, our findings result from age-related differences in disease susceptibility and contact structure. Despite high numbers of contacts for children of all ages, and in particular in the age group of 10 to 20 years old, closing schools appeared to have much less impact on the effective reproduction number than physical distancing measures outside the school environment. In fact, measures effectively reducing non-school contacts, similar to those measures implemented in response to the first pandemic wave in spring 2020, could have prevented a second wave in autumn without school closures. With an estimated effective reproduction number of 1.3 in August 2020, continuation of school closures would have had much lower effects than measures aiming to reduce non-school related contacts, which mainly occur in the adult population. Yet, that situation changes if the proposed measures fail. In November 2020, the measures implemented since August had reduced the effective reproduction number to around 1, instead of achieving the target value of about 0.8. In that situation, as our findings demonstrate, additional physical distancing measures in schools could assist in reducing the effective reproduction number further, in particular when implemented in secondary schools. Our analyses suggest that physical distancing measures in the youngest children will have no impact on the control of SARS-CoV-2 infection. Of note, better adherence to non-school based measures would still have similar effects as reducing school-based contacts.

Although there are several options for reducing the number of contacts between children at school, such as staggered start and end times and breaks, different forms of physical distancing for pupils and division of classes, the effects of such measures on transmission among children have not been quantified. Importantly, we have assumed that reductions in school-based contacts are not replaced by non-school-based contacts with similar transmission risk. Our modelling approach has several limitations. For estimating disease susceptibility we could only model children as group of 0 to 20 years old. As disease susceptibility increases with age, it seems obvious that effects of reduced school contacts are most prominent in older children. Assuming equal susceptibility across these ages may have underestimated to some extent the effect of reducing school contacts for children between 10 and 20 years. At the same time, we assumed that school contact patterns in August-November 2020 reflect the pre-pandemic situation. Yet, universal control measures in the Netherlands such as stay at home orders for symptomatic persons probably lower infectious contacts in school settings too, meaning that some reduction compared to pre-pandemic levels of contacts could already be present in schools. Effects of these measures in school settings should be smaller than in the general population and are hard to estimate due to a large number of asymptomatic cases among children, and therefore were not taken into account. In this respect, the results reported here describe the maximum possible reduction in the effective reproduction number due to school interventions. Furthermore, the contact matrices available did not allow differentiation between various types of contacts outside schools (like work, leisure, transport etc.), as these were not available for periods during the pandemic. Therefore, we could not model the impact of reducing work-related or leisure-related contacts separately. We also could not include hospitalization data from the second wave of the pandemic due to lack of data availability.

The potential effects of opening or closing schools in different phases of the pandemic have been reported in other studies [11, 25–30]. Also based on a mathematical model, Panovska-Griffiths *et al*. [25] predicted that without very high levels of testing and contact tracing reopening schools after summer with a simultaneous relaxation of measures will lead to a second wave in the United Kingdom, peaking in December 2020. Their model predicted that this peak could be postponed for two months (to February 2021) by a rotating timetable in schools. Very early in the pandemic, in March 2020, the Scientific Advisory Group for Emergencies in the United Kingdom, concluded that it would not be possible to get the effective reproduction number below 1 without closing schools [26]. In a modelling study on the impact of non-pharmaceutical interventions for COVID-19 in the United Kingdom, Davies *et al*. found that the impact of school closures was low [11]. In another modeling study Rice *et al*. [30] found that school closures during the first wave of the pandemic could increase overall mortality, due to death being postponed to a second wave. And based on an analysis of the impact of non-pharmaceutical measures in 41 countries between January and May 2020, Brauner *et al*. [27] concluded that closure of schools and universities had contributed the most to lowering the effective reproduction number. Yet, a major difficulty in estimating the effect of school closure based on observational data from the first wave is that other non-pharmaceutical interventions were implemented at or around the same time as school closures [31]. Similarly, lifting such measures often coincided with school re-openings. Observational data from the period after the first wave show conflicting results on within school transmission [32–35] and the effect of school reopening and interpretation is further hampered by the variety in control measures implemented in schools across countries. Finally, Munday *et al*. showed that reopening secondary schools is likely to have a greater impact on community transmission than reopening primary schools in England [28]. While the modelling approach of [28] is different from ours, our findings are similar in the sense that secondary schools are predicted to make a larger contribution to transmission than primary schools, and are therefore more important for controlling COVID-19.

In conclusion, we have demonstrated that the potential effects of school-based measures to reduce contacts between children, including school closures, markedly depends on the reduction in the effective reproduction number achieved by other measures. With remaining opportunities to reduce the effective reproduction number with non-school-based measures, the additional benefit of school-based measures is low. Yet, if opportunities to reduce the effective reproduction number with non-school-based measures are considered to be exhausted or undesired for economic reasons and *R*_*e*_ is still close to 1, the additional benefit of school-based measures may be considerable. In such situations, the biggest impact on transmission is achieved by reducing contacts in secondary schools.

## Data Availability

The data, the R, Stan and Mathematica codes reproducing this study are available at https://github.com/lynxgav/COVID19-schools

## Acknowledgements

The contribution of CHvD was under the auspices of the US Department of Energy (contract number 89233218CNA000001) and supported by the National Institutes of Health (grant number R01-OD011095). MEK was supported by ZonMw grant number 10430022010001, ZonMw grant number 91216062, and H2020 project 101003480 (CORESMA). MJMB and PBV were supported by H2020 project 101003589 (RECOVER). GR was supported by FCT project 131 596787873. We thank Michiel van Boven from the National Institute of Public Health and the Environment, Bilthoven, The Netherlands, for valuable discussions and continuing advice during the course of this project. We thank Mui Pham and Alexandra Teslya for comments on the manuscript.

---

**Figure S1.**
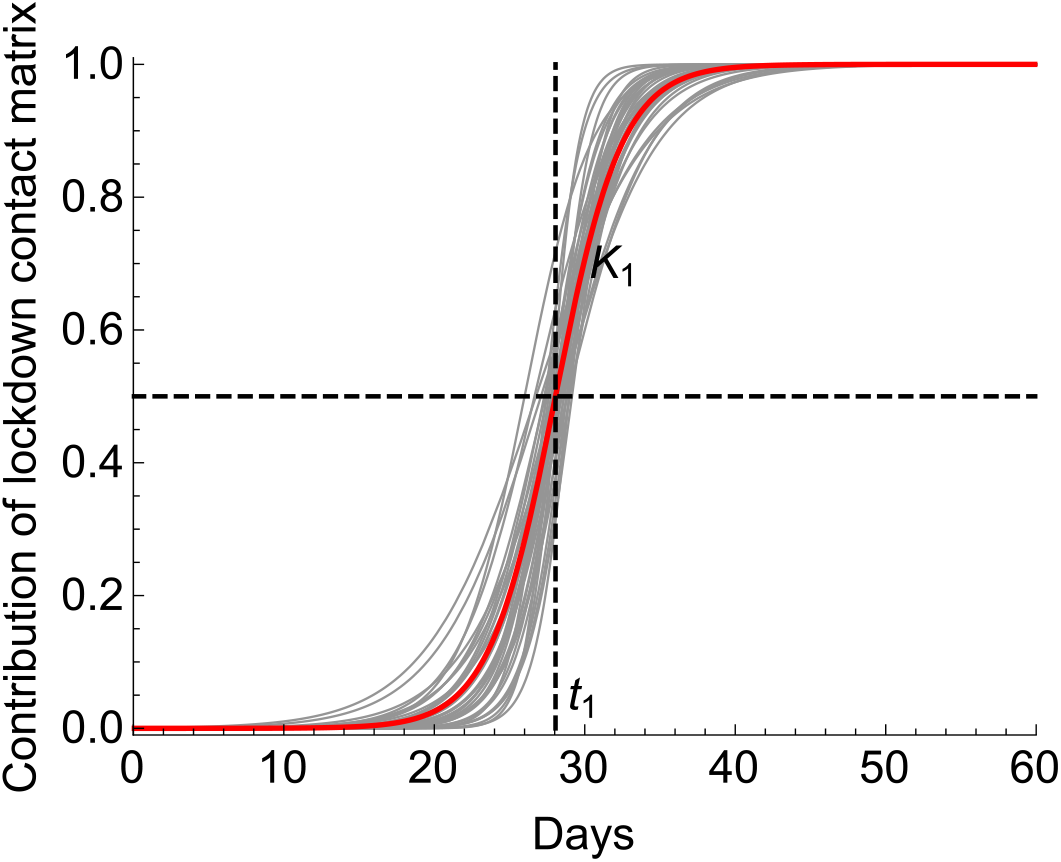
Contribution of the contact rate after the first lockdown. We model the transition in the general contact rate, *c*_*kl*_, as follows *c*_*kl*_ = [1 − *f* (*t*)]*b*_*kl*_ + *ζ*_1_*f* (*t*)*a*_*kl*_, where *f* is the contribution of the contact rate after the first lockdown, *b*_*kl*_ and *a*_*kl*_ are the contact rates specific to the periods before and after the first lockdown. *f* is a logistic function with parameters *K*_1_ and *t*_1_ governing the speed and mid-way of lockdown roll-out. The red and gray lines show the median and several individual estimated trajectories, respectively.

**Figure S2.**
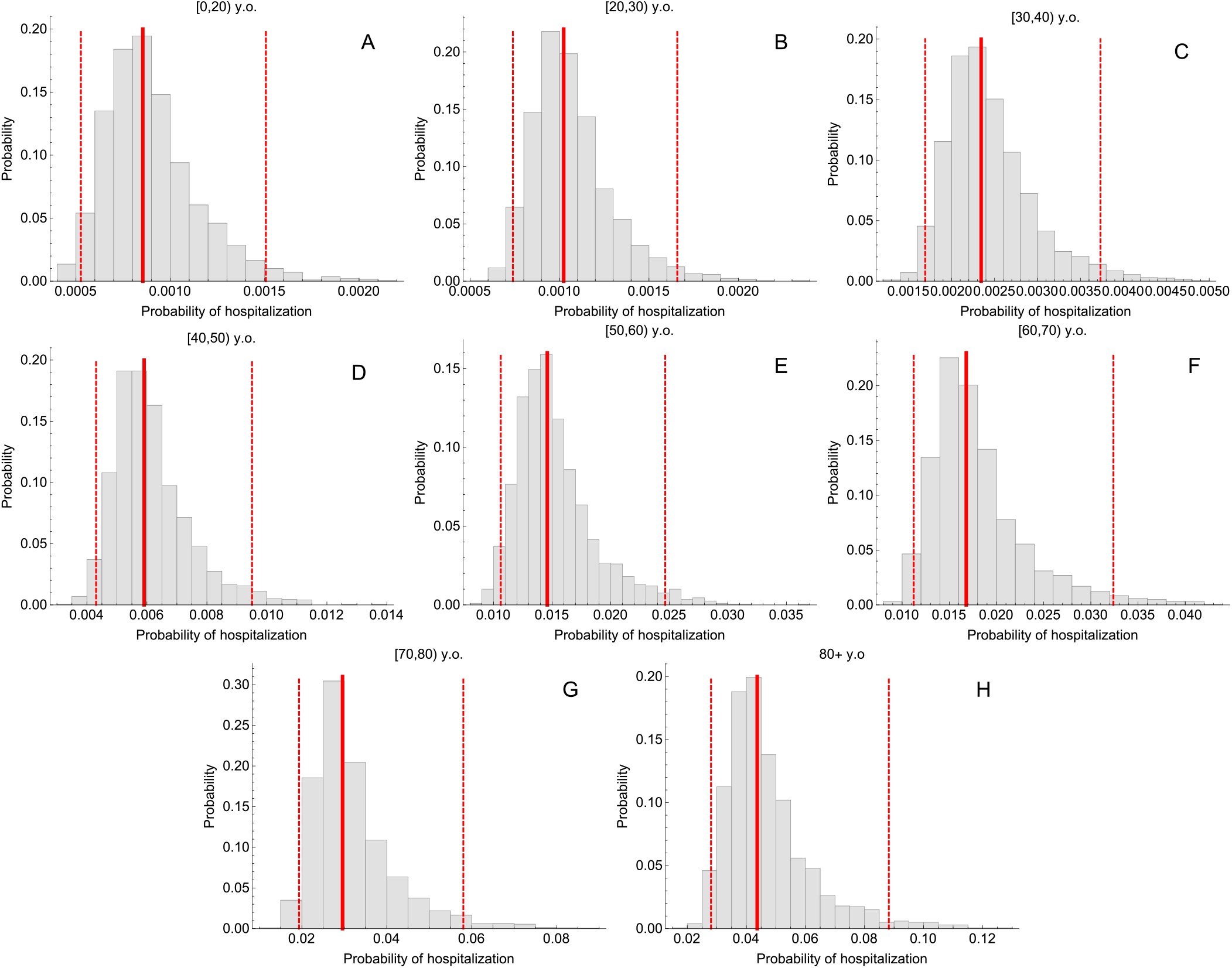
Estimates of probabilities of hospitalization. Histograms are based on 2000 parameter samples from the posterior distribution. The solid and the dashed lines correspond to the median and 95% credible intervals.

**Figure S3.**
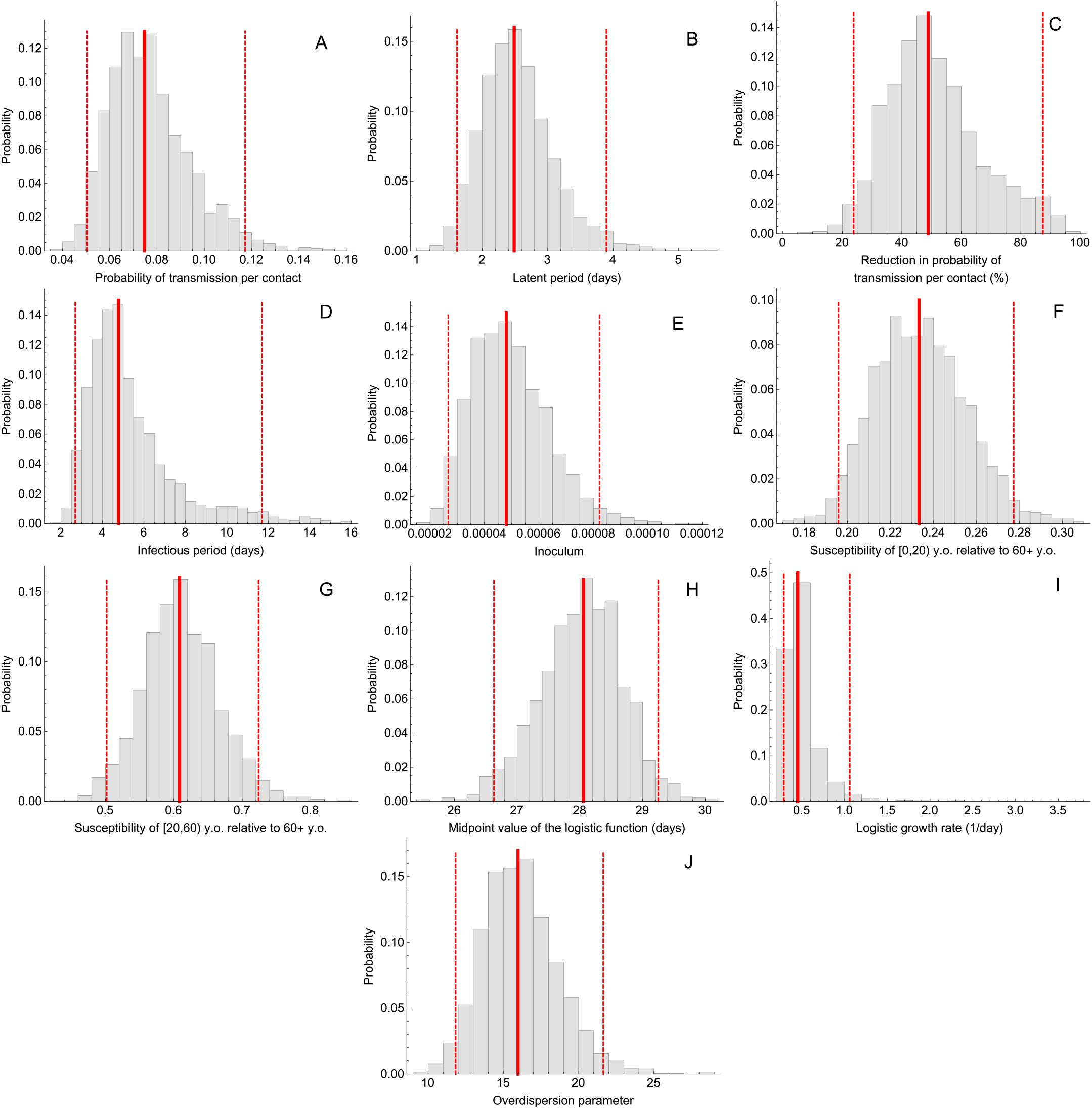
Parameter estimates. Histograms are based on 2000 parameter samples from the posterior distribution. The solid and the dashed lines correspond to the median and 95% credible intervals.

**Figure S4.**
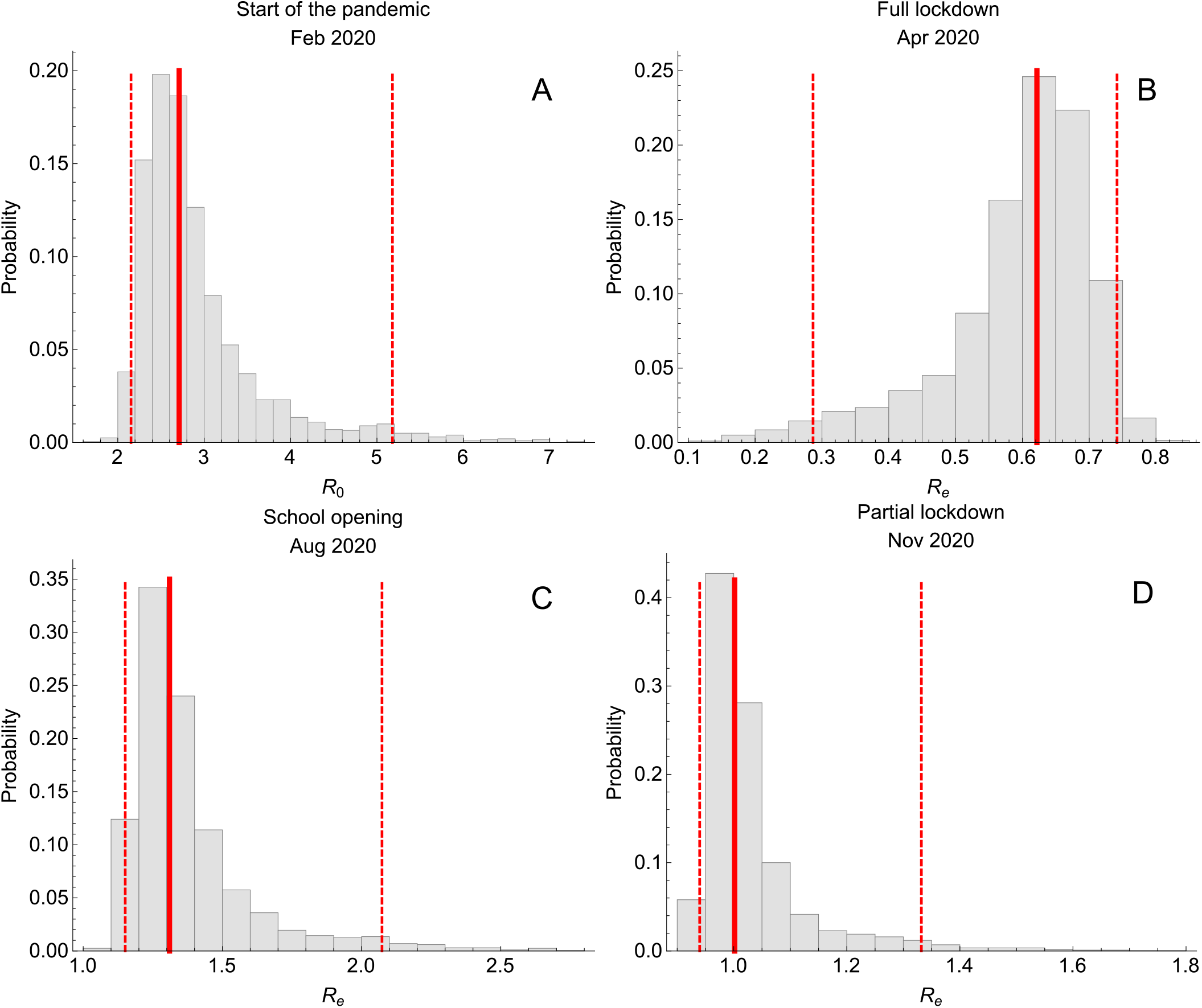
Reproduction numbers. Estimated reproduction numbers (A) at the beginning of the pandemic (February 2020), (B) after the first full lockdown (April 2020), (C) at the time of school opening (August 2020), and (D) after the second partial lockdown (November 2020). Histograms are based on 2000 parameter samples from the posterior distribution. The solid and the dashed lines correspond to the median and 95% credible intervals.

## References

[1] Coronavirus dashboard; 2020. Available from: https://coronadashboard.government.nl/.

[2] Thompson RN, Hollingsworth TD, Isham V, Arribas-Bel D, Ashby B, Britton T, et al. Key questions for modelling COVID-19 exit strategies. Proceedings of the Royal Society B. 2020;287(1932):20201405.

[3] Ferguson NM, Cummings DA, Fraser C, Cajka JC, Cooley PC, Burke DS. Strategies for mitigating an influenza pandemic. Nature. 2006;442(7101):448–452.

[4] Cauchemez S, Ferguson NM, Wachtel C, Tegnell A, Saour G, Duncan B, et al. Closure of schools during an influenza pandemic. The Lancet Infectious Diseases. 2009;9:473–481.

[5] te Beest DE, Birrell PJ, Wallinga J, De Angelis D, van Boven M. Joint modelling of serological and hospitalization data reveals that high levels of pre-existing immunity and school holidays shaped the influenza A pandemic of 2009 in The Netherlands. Journal of The Royal Society Interface. 2015;12(103):20141244. doi:10.1098/rsif.2014.1244.

[6] Mossong J, Hens N, Jit M, Beutels P, Auranen K, Mikolajczyk R, et al. Social contacts and mixing patterns relevant to the spread of infectious diseases. PLOS Medicine. 2008;5(3):1–1. doi:10.1371/journal.pmed.0050074.

[7] Jing Q, Liu M, Zhang Z, Fang L, Yuan J, Zhang A, et al. Household secondary attack rate of COVID-19 and associated determinants in Guangzhou, China: a retrospective cohort study. Lancet Infect Dis. 2020;20(10):1141–1150. doi:10.1016/S1473-3099(20)30471-0.

[8] Goldstein E, Lipsitch M, Cevik M. On the effect of age on the transmission of SARS-CoV-2 in households, schools and the community. The Journal of Infectious Diseases. 2020;doi:10.1093/infdis/jiaa691.

[9] Prem K, Liu Y, Russell T, Kucharski A, Eggo R, Davies N, et al. The effect of control strategies to reduce social mixing on outcomes of the COVID-19 epidemic in Wuhan, China: a modelling study. Lancet Public Health. 2020;5(5):e261–e270. doi:10.1016/S2468-2667(20)30073-6.

[10] Teslya A, Pham TM, Godijk NG, Kretzschmar ME, Bootsma MCJ, Rozhnova G. Impact of self-imposed prevention measures and short-term government-imposed social distancing on mitigating and delaying a COVID-19 epidemic: A modelling study. PLOS Medicine. 2020;17(7):1–21. doi:10.1371/journal.pmed.1003166.

[11] Davies NG, Kucharski AJ, Eggo RM, Gimma A, Edmunds WJ, Jombart T, et al. Effects of non-pharmaceutical interventions on COVID-19 cases, deaths, and demand for hospital services in the UK: a modelling study. The Lancet Public Health. 2020;.

[12] Dehning J, Zierenberg J, Spitzner FP, Wibral M, Neto JP, Wilczek M, et al. Inferring change points in the spread of COVID-19 reveals the effectiveness of interventions. Science. 2020;369(6500). doi:10.1126/science.abb9789.

[13] Giordano G, Blanchini F, Bruno R, Colaneri P, Di Filippo A, Di Matteo A, et al. Modelling the COVID-19 epidemic and implementation of population-wide interventions in Italy. Nature Med. 2020;26:855–860. doi:10.1038/s41591-020-0883-7.

[14] Vos ERA, den Hartog G, Schepp RM, Kaaijk P, van Vliet J, Helm K, et al. Nationwide seroprevalence of SARS-CoV-2 and identification of risk factors in the general population of the Netherlands during the first epidemic wave. Journal of Epidemiology & Community Health. 2020;doi:10.1136/jech-2020-215678.

[15] Statistics Netherlands (CBS); 2020. Available from: https://www.cbs.nl.

[16] Backer JA, Mollema L, Vos RAE, Klinkenberg D, van der Klis FRM, de Melker HE, et al. The impact of physical distancing measures against COVID-19 transmission on contacts and mixing patterns in the Netherlands: repeated cross-sectional surveys in 2016/2017, April 2020 and June 2020. medRxiv. 2020;doi:10.1101/2020.05.18.20101501.

[17] Prem K, Cook AR, Jit M. Projecting social contact matrices in 152 countries using contact surveys and demographic data. PLOS Computational Biology. 2017;13(9):1–21. doi:10.1371/journal.pcbi.1005697.

[18] Diekmann O, Heesterbeek H, Britton T. Mathematical Tools for Understanding Infectious Disease Dynamics. Princeton University Press; 2013.

[19] van Boven M, Teirlinck AC, Meijer A, Hooiveld M, van Dorp CH, Reeves RM, et al. Estimating Transmission Parameters for Respiratory Syncytial Virus and Predicting the Impact of Maternal and Pediatric Vaccination. J Infect Dis. 2020;222(Supplement 7):S688–S694.

[20] Rozhnova G, Kretzschmar ME, van der Klis F, van Baarle D, Korndewal M, Vossen AC, et al. Short- and long-term impact of vaccination against cytomegalovirus: a modeling study. BMC Med. 2020;18. doi: https://doi.org/10.1186/s12916-020-01629-3.

[21] Carpenter B, Gelman A, Hoffman M, Lee D, Goodrich B, Betancourt M, et al. Stan: A probabilistic programming language. J Stat Softw. 2017;76(1):1–32. doi:10.18637/jss.v076.i01.

[22] Diekmann O, Heesterbeek JAP, Roberts MG. The construction of next-generation matrices for compartmental epidemic models. Journal of The Royal Society Interface. 2010;7(47):873–885. doi:10.1098/rsif.2009.0386.

[23] Sekine T, Perez-Potti A, Rivera-Ballesteros O, Strlin K, Gorin JB, Olsson A, et al. Robust T Cell Immunity in Convalescent Individuals with Asymptomatic or Mild COVID-19. Cell. 2020;183(1):158–168.e14. doi: https://doi.org/10.1016/j.cell.2020.08.017.

[24] Burgess S, Ponsford MJ, Gill D. Are we underestimating seroprevalence of SARS-CoV-2? BMJ. 2020;370. doi:10.1136/bmj.m3364.

[25] Panovska-Griffiths J, Kerr C, Stuart R, Mistry D, Klein D, Viner R, et al. Determining the optimal strategy for reopening schools, the impact of test and trace interventions, and the risk of occurrence of a second COVID-19 epidemic wave in the UK: a modelling study. Lancet Child Adolesc Health. 2020;4(11):817–827. doi:10.1016/S2352-4642(20)30250-9.

[26] Scientific Advisory Group for Emergencies. Timing of the introduction of school closure for COVID-19 epidemic suppression, 18 March 2020; 2020. Available from: https://www.gov.uk/government/publications/timing-of-the-introduction-of-school-closure-for-covid-19-epidemic-suppression-18-march-2020.

[27] Brauner JM, Mindermann S, Sharma M, Johnston D, Salvatier J, Gavenčiak T, et al. The effectiveness of eight nonpharmaceutical interventions against COVID-19 in 41 countries. medRxiv. 2020;doi:10.1101/2020.05.28.20116129.

[28] Munday JD, Sherratt K, Meakin S, Endo A, Pearson CAB, Hellewell J, et al. Implications of the school-household network structure on SARS-CoV-2 transmission under different school reopening strategies in England. medRxiv. 2020;doi:10.1101/2020.08.21.20167965.

[29] Keskinocak P, Asplund J, Serban N, Oruc Aglar BE. Evaluating Scenarios for School Reopening under COVID19. medRxiv. 2020;doi:10.1101/2020.07.22.20160036.

[30] Rice K, Wynne B, Martin V, Ackland GJ. Effect of school closures on mortality from coronavirus disease 2019: old and new predictions. BMJ. 2020;371. doi:10.1136/bmj.m3588.

[31] Li Y, Campbell H, Kulkarni D, Harpur A, Nundy M, Wang X, et al. The temporal association of introducing and lifting non-pharmaceutical interventions with the time-varying reproduction number (R) of SARS-CoV-2: a modelling study across 131 countries. The Lancet Infectious Diseases. 2020;doi:https://doi.org/10.1016/S1473-3099(20)30785-4.

[32] Heavey L, Casey G, Kelly C, Kelly D, McDarby G. No evidence of secondary transmission of COVID-19 from children attending school in Ireland, 2020. Eurosurveillance. 2020;25(21). doi:https://doi.org/10.2807/1560-7917.ES.2020.25.21.2000903.

[33] Yung CF, Kam Kq, Nadua KD, Chong CY, Tan NWH, Li J, et al. Novel Coronavirus 2019 Transmission Risk in Educational Settings. Clinical Infectious Diseases. 2020;doi:10.1093/cid/ciaa794.

[34] Macartney K, Quinn H, Pillsbury A, Koirala A, Deng L, Winkler N, et al. Transmission of SARS-CoV-2 in Australian educational settings: a prospective cohort study. Lancet Child Adolesc Health. 2020;4(11):807–816. doi:10.1016/S2352-4642(20)30251-0.

[35] Ismail SA, Saliba V, Lopez Bernal JA, Ramsay ME, Ladhani SN. SARS-CoV-2 infection and transmission in educational settings: cross-sectional analysis of clusters and outbreaks in England. medRxiv. 2020;doi:10.1101/2020.08.21.20178574.

